# Characterisation of Acute Kidney Injury in Critically Ill Patients with Severe Coronavirus Disease-2019 (COVID-19)

**DOI:** 10.1101/2020.05.06.20069872

**Authors:** Sébastien Rubin, Arthur Orieux, Renaud Prevel, Antoine Garric, Marie-Lise Bats, Sandrine Dabernat, Fabrice Camou, Olivier Guisset, Nahema Issa, Gaelle Mourissoux, Antoine Dewitte, Olivier Joannes-Boyau, Catherine Fleureau, Hadrien Rozé, Cédric Carrié, Laurent Petit, Benjamin Clouzeau, Charline Sazio, Hoang-Nam Bui, Odile Pillet, Claire Rigothier, Frederic Vargas, Christian Combe, Didier Gruson, Alexandre Boyer

## Abstract

**Background:** COVID-19-associated acute kidney injury frequency, severity and characterisation in critically ill patients has not been reported.

**Methods:** Single-center cohort performed from March 3, 2020, to April 14, 2020 in 4 intensive care units in Bordeaux University Hospital, France. All patients with COVID19 and pulmonary severity criteria were included. AKI was defined using KDIGO criteria. A systematic urinary analysis was performed. The incidence, severity, clinical presentation, biological characterisation (transient vs. persistent acute kidney injury; proteinuria, hematuria and glycosuria), and short-term outcomes was evaluated.

**Results:** 71 patients were included, with basal serum creatinine of 69 ± 21 µmol/L. At admission, AKI was present in 8/71 (11%) patients. Median follow-up was 17 [12–23] days. AKI developed in a total of 57/71 (80%) patients with 35% Stage 1, 35% Stage 2, and 30% Stage 3 acute kidney injury; 10/57 (18%) required renal replacement therapy. Transient AKI was present in only 4/55 (7%) patients and persistent AKI was observed in 51/55 (93%). Patients with persistent AKI developed a median urine protein/creatinine of 82 [54–140] (mg/mmol) with an albuminuria/proteinuria ratio of 0.23 ± 20 indicating predominant tubulo-interstitial injury. Only 2 (4%) patients had glycosuria. At Day 7 onset of after AKI, six (11%) patients remained dependent on renal replacement therapy, nine (16%) had SCr > 200 µmol/L, and four (7%) died. Day 7 and day 14 renal recovery occurred in 28% and 52 % respectively.

**Conclusion:** COVID-19-associated AKI is frequent, persistent severe and characterised by an almost exclusive tubulo-interstitial injury without glycosuria.

## Introduction

Since December 2019, COVID-19 has infected nearly two million people worldwide, leading to more than 120 000 deaths. Starting in China, it rapidly expanded in Europe, affecting first Italy, then Spain and France. Globally, it has resulted in the confinement of nearly three billion people ^1,2^. The severe respiratory damages requiring prolonged intubation have already been described. They are the determinant of intensive care unit (ICU) admission and have resulted in a consecutive risk of ICU saturation^3–7^.

Acute kidney injury (AKI) is the second most frequent organ damage previously reported in China. It is observed in up to 15% of patients, whatever their need for ICU admission^4^. However, very few data are available in patients admitted to ICUs, where this incidence should be significantly higher. According to the first United States report of ICU patients (n = 24) ^6^, it is observed in 20% of subjects; this does not reflect the first Western European findings where more than 50% of patients could experience AKI (unpublished data). The reason for this is not clearly established. AKI severity and potential recovery have not been reported. The need for renal replacement therapy (RRT) may considerably prolong the length of ICU hospitalization, leading to significant organizational and economic challenges for health care systems. In addition, a high frequency of AKI could result in a very significant increase in the number of patients suffering from long-term chronic kidney disease (CKD) ^8^. The hypothesis is that COVID-19-associated AKI is more severe and common than previously reported and may be represented by different clinical presentations including transient AKI, acute tubular necrosis, or other types of injury. Each of these clinical presentations potentially requires a specific approach. The objective of the study is to describe the incidence and severity and to characterise COVID-19-associated AKI.

## Materials and methods

### Study design

This cohort study was carried out in the four ICUs dedicated to severe COVID-19 patients at the University Hospital of Bordeaux, France, from March 3, 2020, to April 14, 2020. Patients’ data were routinely collected during their entire hospital stay in dedicated electronic health records. According to French law and the French Data Protection Authority, the handling of these data for research purposes was declared to the Data Protection Officer of the University Hospital of Bordeaux. The study obtained the approval of the Institutional Review Board of the University Hospital of Bordeaux (declaration number 2020–14). Patients (or their relatives, if any) were notified about the anonymized use of their healthcare data via the department’s booklet.

### Participants

Patients were enrolled if they were aged 18 years or older, had a positive polymerase chain reaction for COVID-19 or typical computed tomography findings in patients with a high clinical pre-test probability of COVID-19^9^ and had severity criteria for admission to one of four ICUs. These severity criteria were defined by the need of ≥ 4 L/min oxygen therapy (O_2_) to obtain an arterial oxygen saturation (SaO_2_) ≥ 94% or arterial p artial p ressure of oxygen/fraction of inspired oxygen ratio (PaO_2_/FiO_2_) ≦ 300 mmHg in patients receiving high-flow nasal cannula oxygen, non-invasive or invasive mechanical ventilation. The University Hospital of Bordeaux was confronted with a far less aggressive epidemic context than those reported in other French regions or other countries. Therefore, all patients presenting with this severity were systematically admitted to ICU except those who had withdrawal orders. No specific withdrawal policy was applied during this epidemic period. ICU patients were excluded from this study only if they had pre-existing end-stage kidney disease defined by the need for Renal Replacement Therapy.

## Patients management

Patients were treated according to standard of care. In three units, hydroxychloroquine (HCL) was administered because of physicians’ intimate decision according to physiopathological studies suggesting an in vitro efficacy against SARS-CoV-2. The dosage schedule was per os 400mg b.i.d. at day 1 and then 200mg b.i.d. for 5 to 10 days. One of this unit added tocilizumab to HCL with a single i.v. 8mg/kg infusion. Two units administered lopinavir/ritonavir (400mg b.i.d. syrup form) at first place and then stopped because of drug shortage. All treatments were started the day of admission to ICU Importantly, no other modification in COVID-19 critically ill patients care occurred and treatment administration were dependent on the unit and not on patients’ characteristics. All patients had foley catheterization.

### Acute kidney injury classification

The KDIGO definition was used to define AKI Stages 1, 2, and 3^10^. Serum creatinine (SCr) and urine output were both taken into account. Stage 1: SCr 1.5–1.9 times baseline or diuresis < 0.5 mL/kg/h for 6–12 hours; Stage 2: SCr 2.0–2.9 times baseline or diuresis < 0.5 mL/kg/h for ≥ 12 hours; Stage 3: SCr ≥ 3 times baseline, initiation of renal replacement therapy, anuria ≥ 12 hours, or diuresis < 0.3 mL/kg/h for ≥ 24 hours. AKI stage was classified using the worst SCr or diuresis during the entire ICU stay.

Baseline SCr values corresponded to SCr values at admission in the case of normal renal function or SCr values from within six months in the case of abnormal SCr at admission. All SCr baseline values could be collected from previous medical records for patients with abnormal SCr at ICU admission.

Acute kidney disease (AKD) staging was performed using consensus report of the Acute Disease Quality Initiative (ADQI) 16 workgroup^10^ and was defined as a condition wherein criteria for AKI stage 1 or greater persists ≥7 days after an exposure. AKD stage 1: SCr 1.5-1.9 times baseline; Stage 2: SCr 2.0–2.9 times baseline, Stage 3: SCr ≥ 3 times baseline or increase in serum creatinine ≥ 353.6µmol/L or ongoing need for renal replacement therapy.

### Exposure variables

Information relative to drugs taken before admission, date and type of the first COVID-19 symptoms, and a medical history of hypertension, diabetes, chronic heart failure (chronic ischemic, rhythmic, valvular heart disease or heart failure with preserved ejection fraction or reduced ejection fraction), stroke, chronic obstructive pulmonary disease, asthma and peripheral arterial disease were collected using prospectively recorded data and patient or relative questioning. Pre-admission exposure to nonsteroidal anti-inflammatory drugs and renin-angiotensin system inhibitors was considered if the patient had taken the drug the week prior to admission. The reason for ICU admission, weight, and height were measured at ICU admission. All other healthcare data were collected using Metavision ICU (iMDsoft). The four ICUs of the University Hospital of Bordeaux received patients (n = 16) transferred from the ICUs of the Mulhouse or Strasbourg Hospitals in France. They were included whenever they met the inclusion criteria of the ICU admission cohort. Healthcare data were collected using medical records from the previous hospital.

Patients chronically exposed to immunosuppressive drugs or suffering from hematologic malignancies were considered immunosuppressed.

CKD was defined as an estimated glomerular filtration rate of < 60 mL/min/1.73m2, using the Chronic Kidney Disease Epidemiology Collaboration corresponding to CKD Stage 3 or more according to the KDIGO classification^10^.

Fluid balance was the addition of fluid infusion volume, including enteral or parenteral nutrition minus diuresis and gastrointestinal losses.

Minimum diuresis was defined by the minimum diuresis/kg/hour used for the KDIGO classification.

All blood or urine biological assessments were usually standardized in ICUs participating in the study and systematically collected. All biochemical parameters were determined on Abbot Architect analyzers with enzymatic method for creatinine and colorimetric bromocresol purple method for albumin. SCr was measured at least once a day. A urinary analysis was performed at ICU admission or whenever AKI occurred or deteriorated during the ICU stay. Fractional excretion of sodium (FeNa^+^) and fractional excretion of urea (FeUrea) was calculated as follows: FeNa^+^ (%) = ([U/P sodium] / [U/P creatinine]) x 100; FeUrea (%) = ([U/P urea] / [U/P creatinine]) x 100. U/P is urinary/plasma ratio, and urinary osmolarity was measured.

The AKI day was the first day during which the patient was eligible for AKI using KDIGO classification.

### Outcomes definitions

Since 2017, RRT initiation criteria have been standardized in the ICUs participating in the study, according to the delayed strategy of the Artificial Kidney Initiation in Kidney Injury (AKIKI) trial^11^. Patients were eligible for RRT whenever AKI Stage 3 occurred with clinical indications (anuria ≥ 72hours, [K+] > 6 mmol/L or > 5.5 mmol/L after medical correction, acute pulmonary edema, pH < 7.15 in the absence of other causes, or blood urea > 40 mmol/L).

The ICU length of stay was only calculated for patients that were discharged from the ICU; most of them were transferred to medical wards or a continuing care unit.

The intubation length was only calculated for the subgroup of patients who were successfully extubated. If a patient ever needed to be reintubated after having been weaned from the ventilator, the intubation length was calculated as the sum of the different periods during which invasive mechanical ventilation was required.

Transient AKI was defined by a ≥ 50% decrease in SCr or a return of SCr to its baseline value 72h after AKI^12^.

Recovery from AKI was defined as a return of SCr to ≤ 125% above baseline SCr for alive and non-dependent RRT patients during the ICU stay.^13^

Major Adverse Kidney Events (MAKE) was a composite outcome defined by an SCr > 200% of baseline, need for RRT, or death^14^.

### Statistical analysis

Statistical analysis was carried out using JMP^®^ Version 14, SAS Institute Inc., Cary, NC, 1989–2007. Descriptive statistics included mean ± standard deviation or median [Quartile 1–Quartile 3] if the variable did not fit a normal distribution. Quantitative variables were compared using a t-test, and qualitative variables were compared using Fisher’s exact test when only two variables were studied; Pearson’s Chi-squared test was used for more variables. AKI incidence during ICU hospitalization and renal recovery during hospitalization was plotted using a Kaplan–Meier curve with a 95% confidence interval and was censured with death. A value of double-sided p < 0.05 was considered statistically significant.

## Results

From March 3, 2020, 380 patients were admitted in the University Hospital of Bordeaux for a COVID-19. Among them, 45/380 (12%) patients with severe COVID-19 were admitted to the ICU in addition to 26 critically ill patients admitted to ICU from other regions of France for a total of 71 patients included (figure 1). The reason for admission was acute respiratory failure for 70/71 (99%), with a majority of males – 55/71 (77%) – and a mean age of 61 ± 12 years. Time from the onset of first viral symptoms to ICU admission was eight [6–10] days. Details on all baseline characteristics, including blood test results at admission, are presented in Table 1.

**Table 1.**
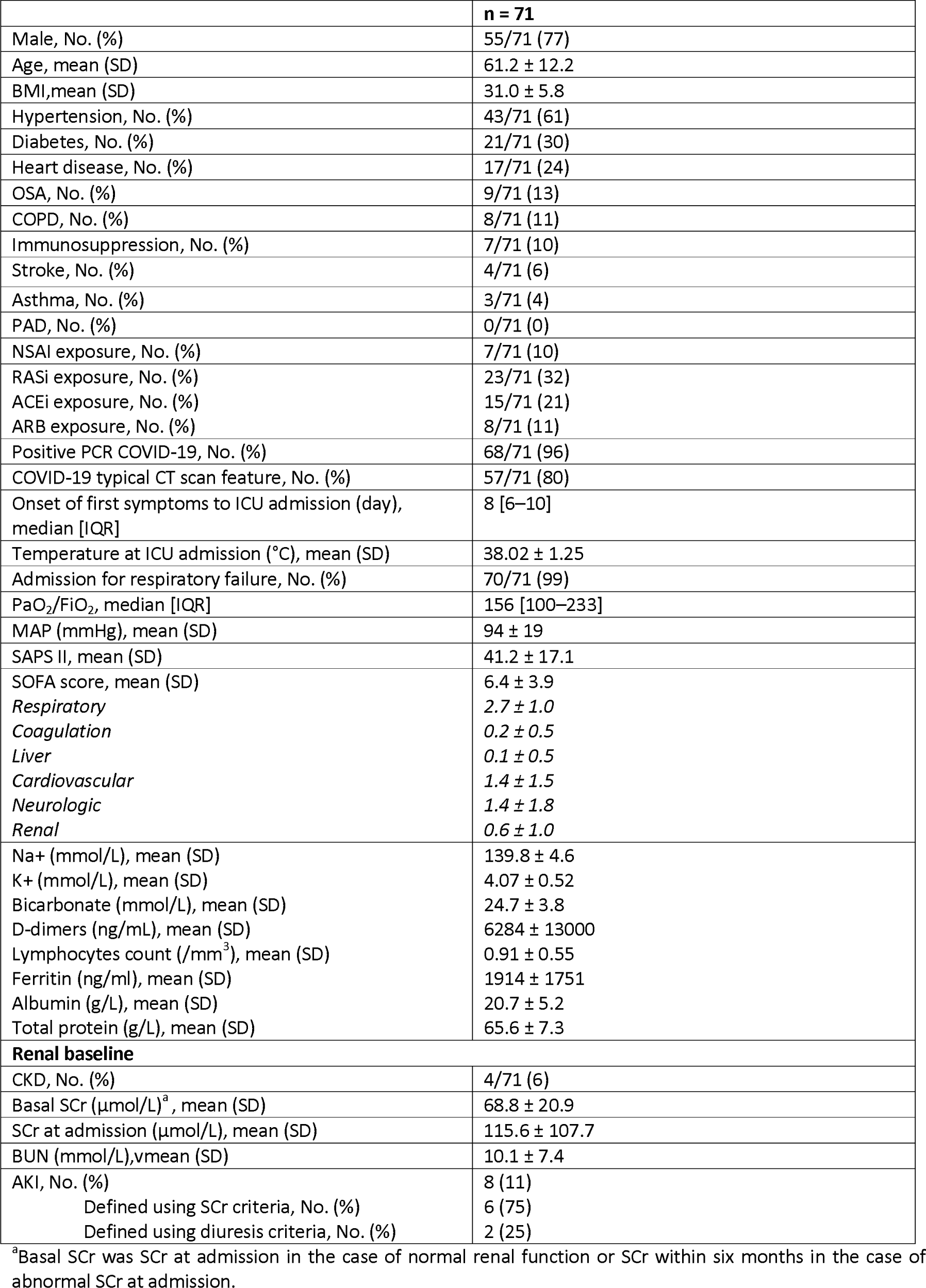
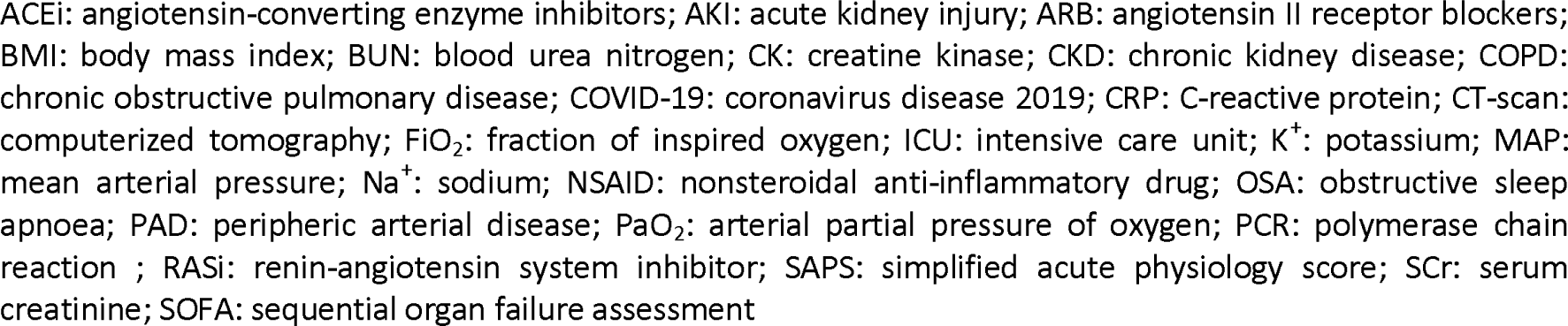
Baseline characteristics of patients at ICU admission.

|  | **n = 71** |
| --- | --- |
| Male, No. (%) | 55/71 (77) |
| Age, mean (SD) | 61.2 ± 12.2 |
| BMI, mean (SD) | 31.0 ± 5.8 |
| Hypertension, No. (%) | 43/71 (61) |
| Diabetes, No. (%) | 21/71 (30) |
| Heart disease, No. (%) | 17/71 (24) |
| OSA, No. (%) | 9/71 (13) |
| COPD, No. (%) | 8/71 (11) |
| Immunosuppression, No. (%) | 7/71 (10) |
| Stroke, No. (%) | 4/71 (6) |
| Asthma, No. (%) | 3/71 (4) |
| PAD, No. (%) | 0/71 (0) |
| NSAI exposure, No. (%) | 7/71 (10) |
| RASi exposure, No. (%) | 23/71 (32) |
| ACEi exposure, No. (%) | 15/71 (21) |
| ARB exposure, No. (%) | 8/71 (11) |
| Positive PCR COVID-19, No. (%) | 68/71 (96) |
| COVID-19 typical CT scan feature, No. (%) | 57/71 (80) |
| Onset of first symptoms to ICU admission (day), median [IQR] | 8 [6–10] |
| Temperature at ICU admission (°C), mean (SD) | 38.02 ± 1.25 |
| Admission for respiratory failure, No. (%) | 70/71 (99) |
| PaO<sub>2</sub>/FiO<sub>2</sub>, median [IQR] | 156 [100–233] |
| MAP (mmHg), mean (SD) | 94 ± 19 |
| SAPS II, mean (SD) | 41.2 ± 17.1 |
| SOFA score, mean (SD) | 6.4 ± 3.9 |
| *Respiratory* | 2.7 ± 1.0 |
| *Coagulation* | 0.2 ± 0.5 |
| *Liver* | 0.1 ± 0.5 |
| *Cardiovascular* | 1.4 ± 1.5 |
| *Neurologic* | 1.4 ± 1.8 |
| *Renal* | 0.6 ± 1.0 |
| Na<sup>+</sup> (mmol/L), mean (SD) | 139.8 ± 4.6 |
| K<sup>+</sup> (mmol/L), mean (SD) | 4.07 ± 0.52 |
| Bicarbonate (mmol/L), mean (SD) | 24.7 ± 3.8 |
| D-dimers (ng/mL), mean (SD) | 6284 ± 13000 |
| Lymphocytes count (/mm<sup>3</sup>), mean (SD) | 0.91 ± 0.55 |
| Ferritin (ng/mL), mean (SD) | 1914 ± 1751 |
| Albumin (g/L), mean (SD) | 20.7 ± 5.2 |
| Total protein (g/L), mean (SD) | 65.6 ± 7.3 |
| **Renal baseline** |  |
| CKD, No. (%) | 4/71 (6) |
| Basal SCr (μmol/L)<sup>a</sup>, mean (SD) | 68.8 ± 20.9 |
| SCr at admission (μmol/L), mean (SD) | 115.6 ± 107.7 |
| BUN (mmol/L), mean (SD) | 10.1 ± 7.4 |
| AKI, No. (%) | 8 (11) |
| Defined using SCr criteria, No. (%) | 6 (75) |
| Defined using diuresis criteria, No. (%) | 2 (25) |

<sup>a</sup>Basal SCr was SCr at admission in the case of normal renal function or SCr within six months in the case of abnormal SCr at admission.
ACEi: angiotensin-converting enzyme inhibitors; AKI: acute kidney injury; ARB: angiotensin II receptor blockers; BMI: body mass index; BUN: blood urea nitrogen; CK: creatine kinase; CKD: chronic kidney disease; COPD: chronic obstructive pulmonary disease; COVID-19: coronavirus disease 2019; CRP: C-reactive protein; CT-scan: computerized tomography; $\text{FiO}_2$ : fraction of inspired oxygen; ICU: intensive care unit; $\text{K}^+$ : potassium; MAP: mean arterial pressure; $\text{Na}^+$ : sodium; NSAID: nonsteroidal anti-inflammatory drug; OSA: obstructive sleep apnoea; PAD: peripheral arterial disease; $\text{PaO}_2$ : arterial partial pressure of oxygen; PCR: polymerase chain reaction ; RASi: renin-angiotensin system inhibitor; SAPS: simplified acute physiology score; SCr: serum creatinine; SOFA: sequential organ failure assessment

**Figure 1.**
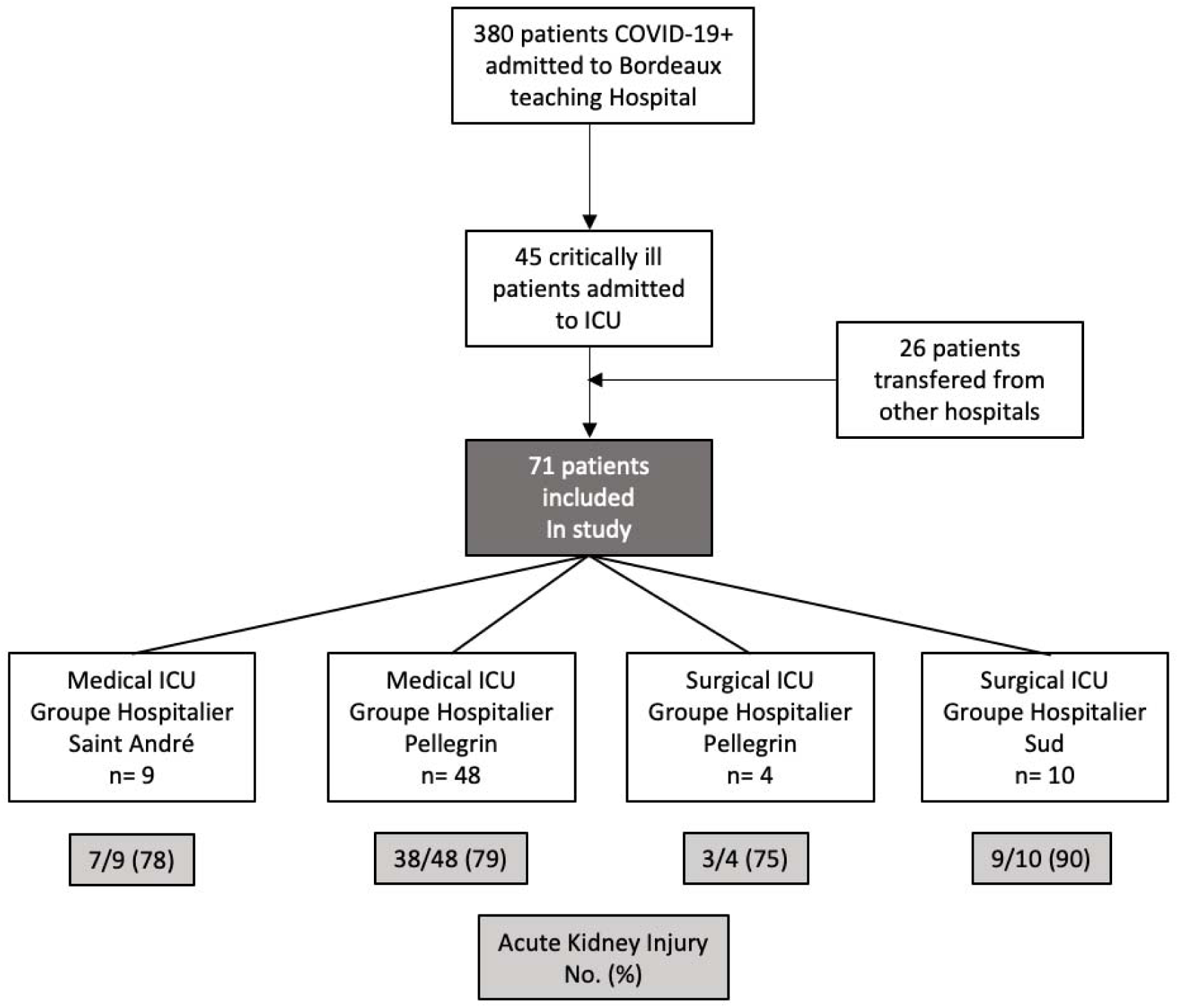
Flow Diagram. ICU: Intensive Care Unit

Median follow-up was 17 [12–23] days, length of ICU stay was 10 [7–14] days. Intubation was required for 55/71 (77%) patients for a median duration of 12 [9–17] days. The worst PaO_2_/FiO_2_ was 95 [79–125]. Septic shock was rare since norepinephrine infusion rate (maximum 0.16 [0.10–0.37] µg/kg/min) was low and no lactate level exceeded 2 mmol/l except in 2 patients^15^. Four patients died (11%). Table 2 shows the main follow-up outcomes.

**Table 2.** Main follow-up outcomes.

|  | **n = 71** |
| --- | --- |
| Follow-up (days), median [IQR] | 17 [12–23] |
| ICU length of stay (n = 38)<sup>a</sup>, median [IQR] | 10 [6.7–14.2] |
| Intubation, No. (%) | 55/71 (77) |
| Length of intubation (n = 29)<sup>b</sup>, median [IQR] | 12 [9–17] |
| Worst PaO<sub>2</sub>/FiO<sub>2</sub>, median [IQR] | 95 [79–125] |
| Need for NE infusion, No. (%) | 52/71 (73) |
| Maximal dose of NE infusion (µg/kg/min), median [IQR] | 0.16 [0.10–0.37] |
| Day of NE maximal dose from ICU admission (days), median [IQR] | 4 [1–12] |
| Maximal lactate level (mmol/L), mean (SD) | 1.5 ± 0.7 |
| ICU deaths, No. (%) | 4/71 (6) |
| Onset of death from ICU admission (days), median [IQR] | 9 [5–19] |

<sup>a</sup> The ICU length of stay was only calculated for patients that were discharged from the ICU, n = 38.
<sup>b</sup> The length of intubation was only calculated for patients that were successfully weaned from the ventilator, n = 29
FiO<sub>2</sub>: fraction of inspired oxygen; ICU: intensive care unit; IQR: interquartile range NE: norepinephrine; PaO<sub>2</sub>: arterial partial pressure of oxygen; SD: standard deviation

### Acute kidney injury

At admission, AKI was present in 8/71 (11%) patients. During their ICU stay, AKI developed in 49 other patients, reaching a total of 57/71 (80%) patients. The timing of AKI development is shown in Figure 2a. Stage 1 AKI was observed in 20/57 (35%) patients, Stage 2 in 20/57 (35%), and Stage 3 in 17/57 (30%); 10/57 (18%) required RRT. Patients with AKI had a higher body mass index (32 ± 6 vs. 28 ± 5; p = 0.04); higher total sequential organ failure assessment (SOFA) (7 ± 3.8 vs. 3.9 ± 3.3; p = 0.006), renal SOFA (0.7 ± 1.1 vs. 0 ± 0; p < 0.001), and cardiovascular SOFA (1.6 ± 1.5 vs. 0.4 ± 0.9, p = 0.004) scores; and lower PaO_2_/FiO_2_ ratios (93 [76–120] vs. 109 [90–227]; p = 0.04). They had similar maximal norepinephrine infusion (0.16 [0.10–0.39] vs. 0.23 [0.12-0.30]; p = 0.76), maximal lactate level (1.4 ± 0.7 mmol/L vs. 1.5 ± 0.3 mmol/L; p = 0.76), contrast agent infusion (12/57 (21%) vs. 3/14 (21%); p = 0.98), and crystalloid infusion during the first 24h after admission (3074 ± 1577 ml. vs. 2761 ± 905 ml.; p = 0.37). Comparisons between patients with or without AKI is presented in Table 3.

**Table 3.**
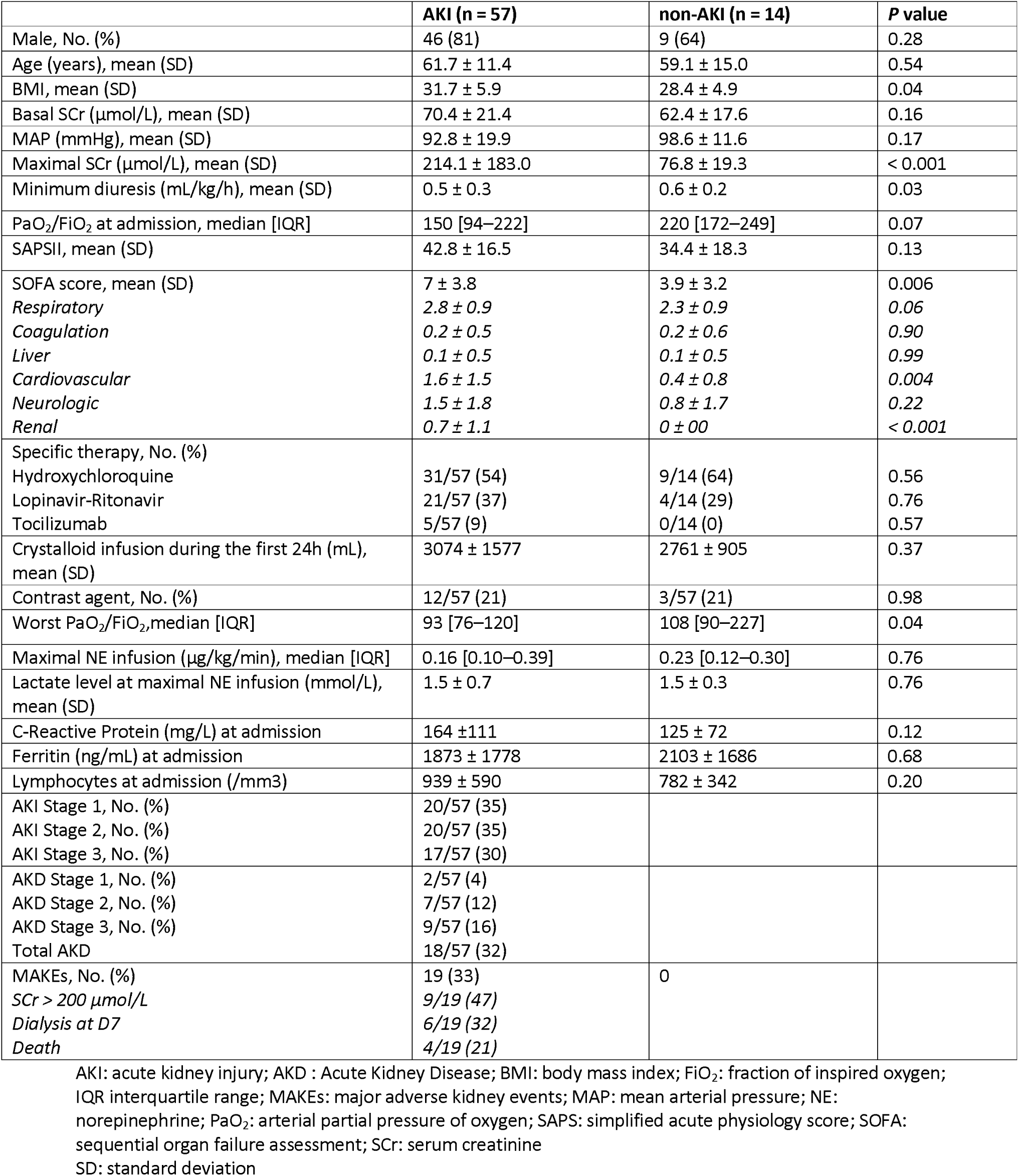
Comparison of patients with and without AKI.

**Figure 2.**
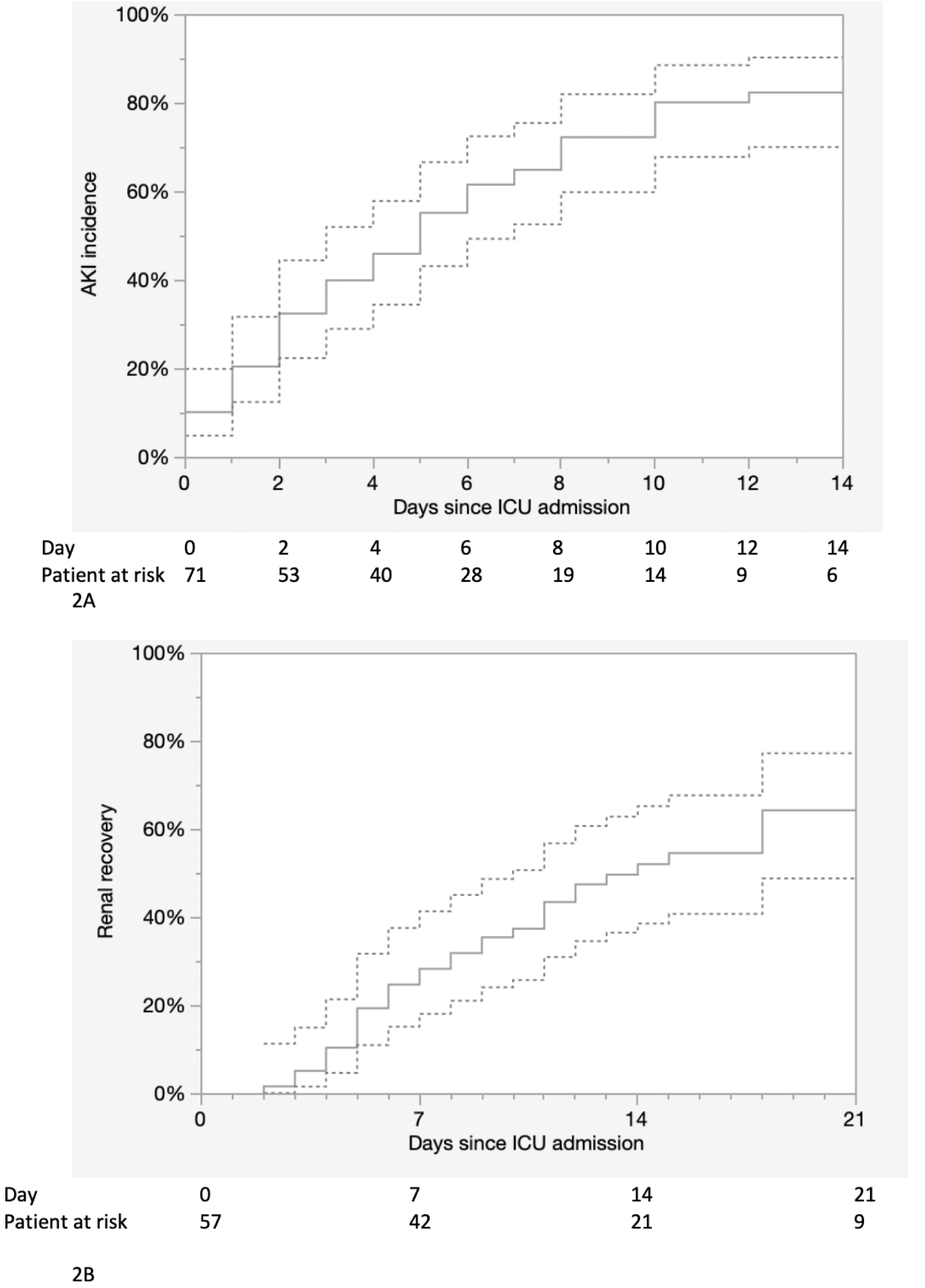
2a: Incidence of AKI during the ICU stay censored with death with 95 % confident interval. 2b: Renal recovery during hospitalization censored with death with 95% confident interval.

### Characterisation of acute kidney injury

Because of two deaths in the first 72h after AKI, 55 patients could be assessed to differentiate transient from persistent AKI. Transient AKI was present in only 4/55 (7%) patients, and persistent AKI was present in 51/55 (93%). Compared to patients with persistent AKI, patients with transient AKI had similar basal SCr (69 ± 21 µmol/L vs. 76 ± 23 µmom/L; p = 0.62), natriuresis (53 ± 40 mmol/L vs. 81 ± 75 mmol/L; p = 0.52), urinary osmolarity (515 ± 182 mmol/L vs. 637 ± 330 mmol/L; p = 0.52), FeNa^+^ (0.6 ± 0.9% vs. 0.6 ± 0.7%; p = 0.96), FeUrea (33 ± 12% vs. 39 ± 11%; p = 0.35), and fluid balance at 72h after AKI diagnosis (12805 ± 6378 mL vs. 8070 ± 3732 mL; p = 0.29), respectively (Table 4).

**Table 4.**
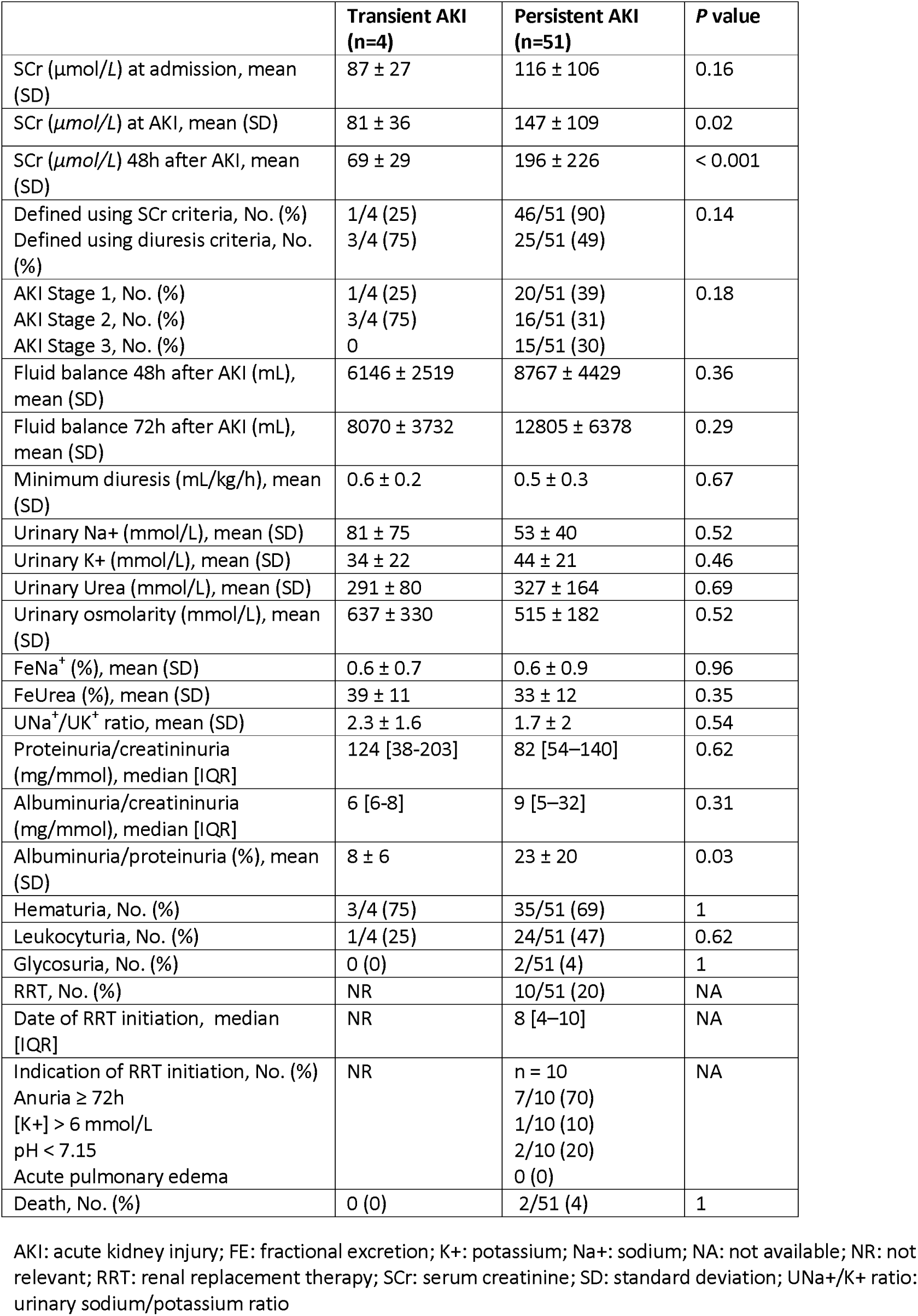
Comparison between patients with transient vs. persistent AKI.

Patients with persistent AKI developed a median proteinuria/creatininuria of 82 [54–140] mg/mmol with an albuminuria/proteinuria ratio of 0.13 [0.07-0.32], Hematuria was present in 35/51 (69%) patients; leukocyturia was present in 24/51 (47%) patients. Glycosuria was found in 2/51 (4%) patients.

### Renal outcomes

At Day 7 after AKI, 6/57 (11%) patients remained dependent on RRT, 9/57 (16%) had SCr > 200 µmol/L, and 4/57 (7%) died, accounting for 19/57 (33%) patients reaching MAKE criteria. Renal recovery occurred in 28% of patients at Day 7, 52% at Day 14, and 64% at Day 21 (Figure 2b).

## Discussion

In this study, the incidence of COVID19-associated AKI was higher (80%) than reported previously (3-20%)^4,7,16^. Cases was severe, with 30% stage 3 and 18% RRT and not transient - more than 90% of cases were persistent AKI. The value of urinary indices to differentiate transient vs. persistent AKI in this context might be questioned. Both high proteinuria and low albuminuria were not in favor of a prevalent glomerular injury and suggest acute tubular injury (ATI), interstitial nephritis (IN) or both. The very low incidence of glycosuria was not in favor of Fanconi syndrome. Hematuria and leukocyturia could not be interpreted by the presence of foley catheter for all patients.

To the best of our knowledge, this is the fisrt study to systematically explore COVID19-associated AKI, particularly in more severe patients. The higher incidence of AKI observed in this study can be explained in different ways. First, it could be related to the severity of the disease by an increase in the initial renal viral charge and/or severe systemic inflammation. Moreover, the exhaustive collection of basal SCr of ICU patients could participate in this higher incidence, providing a more real spectrum of the COVID-19-associated renal injury.

Whether the cause of AKI is ATI, IN or both is debatable. Acute tubular necrosis (ATN), the most common form of ATI, is mainly influenced by to hemodynamic parameters and iodinated contrast agents^17^. In this study, the use of these agents was limited (20%). The hemodynamic instability of patients with AKI was slight and due to high sedation infusion rate^18^. No septic shock was observed and no differences in inflammatory states (C-Reactive Protein and Ferritin) was observed between Aki and no AKI patients. This suggests the possibility of a viral specific ATI. SARS-coV-2 (the virus causing COVID19) penetrates the cells via two receptors (ACE2 and TMPRSS2) ^19^. While ACE2 is highly expressed in proximal tubular epithelial cells and in podocytes, TMPRSS2 are only detectable in the proximal tubule S3 segment. By infiltrating the renal tubular cell, SARS-coV-2 might induce ATI. This could explain why 9 out of 26 autopsied patients in China with COVID19-associated AKI had primarily diffuse proximal tubular injury, with some frank necrosis and no glomerular injury^20^. Another postmortem study showed severe ATI with IN (CD68+ macrophage infiltration of the tubulointerstitium)^21^. However, in these post-mortem studies, patients had less severe renal involvement and may be not representative of more severe patients. Pulmonary damages have been reported as an inflammatory pattern integrated into a cytokine storm syndrome^22^. The frequent association we report between renal and pulmonary injuries might have several explanations. First co-occurrence does not necessarily mean a common pathway. However, one is left to wonder if infection with SARS-Cov2 may result in inflammation or direct viral injury of both organs. Renal biopsies in patients with severe COVID19-associated AKI might thus be considered and lead to specific anti-inflammatory therapies for IN-predominant AKI

While FeUrea and FeNa+ have been previously reported as relevant for the differentiation of transient-AKI from persistent AKI^23,12^ this was not shown in the current study.

Limitations of our work includes the small number of patients (n=71) and a limited median follow-up period (2 weeks). These results will have to be confirmed by larger studies with longer follow-up period.

In conclusion, this study should make physicians aware of the likely existence of a frequent, severe and specific COVID19-associated AKI. The study provides additional insights into the characterisation of AKI: a tubulo-interstitial injury without glycosuria. Further work should be carried out promptly in order to identify and assess specific therapeutic options.

## Data Availability

N/A

## Acknowledgement

We would like to thank the subjects and the clinical and clerical teams involved in this study. We also thank Philippe Real (MD), Surgical Intensive Care Unit, Mulhouse Hospital; Khaldoun Kuteifan (MD) and Philippe Guiot (MD), Medical Intensive Care Unit, Mulhouse Hospital; Ferhat Meziani (MD), Medical Intensive Care Unit, CHU Strasbourg; Pierre Diemunsch (MD) Surgical Intensive Care Unit, CHU Strasbourg.

